# Greater scaling and less flexible timing of cortical responses during challenging reactive balance after stroke

**DOI:** 10.64898/2026.10.21.26366125

**Authors:** Jacqueline A Palmer, Lena H Ting, Michael R Borich, Aiden M Payne, Jasmine L Mirdamadi

**Author notes:** **Corresponding author:** Dr. Jacqueline A Palmer R219 426 Church St SE Minneapolis, MN 55455. senior co-authors (Aiden M Payne, Jasmine L Mirdamadi).

## Abstract

**Background:** Cortical N1 responses to balance perturbations are larger and earlier with increasing magnitude and differ with reactive stepping. Whether stroke alters this modulation is unknown.

**Objective:** To compare N1 modulation across perturbation magnitudes and reactive balance outcomes in older adults with and without stroke. We hypothesized that people with stroke would show greater N1 scaling than controls; that larger N1s at small perturbations would limit scaling (a recruitment ceiling); and that stroke would constrain N1 modulation by stepping outcome.

**Methods:** Sixteen people with chronic stroke (66±11 years) and 15 age-similar controls (68±8 years) received forward support-surface perturbations at a small standardized magnitude and at individual step thresholds (∼50% unintentional steps). N1 peak amplitude and latency were compared across magnitudes in feet-in-place trials, and between stepping and feet-in-place trials at step threshold. N1 scaling was the relative amplitude increase between magnitudes.

**Results:** Despite lower clinical balance after stroke (p<0.001), step thresholds were similar between groups. N1s were larger with shorter latencies at step threshold in both groups (p<0.001), but scaled more after stroke (44% vs 13%,p=0.009). Greater scaling was associated with smaller small-magnitude N1s after stroke (β=−0.43, p=0.041) and higher step thresholds across groups (β=0.35, p=0.048). N1 latency was 11 ms longer in stepping trials in controls but not after stroke (interaction p=0.024).

**Conclusions:** Scaling of cortical recruitment with increasing perturbation magnitude may support reactive balance, and may be an adaptive mechanism for people after stroke. Less flexible N1 timing after stroke may limit cortical tuning to balance recovery strategy.

## Introduction

Reactive balance impairments in people after stroke limit mobility and independence in daily activities and increasing fall risk.^1,2^ Recovering balance after an unexpected disturbance requires rapid recruitment of both subcortical and cortical resources, with cortical contributions becoming more prominent as balance challenge increases.^3–6^ Lesions affecting cortical and subcortical pathways may therefore compromise the neural resources available for effective balance recovery after stroke.^7^ Consistent with this notion, we previously observed relationships between cortical activity and balance behavior after stroke under conditions that preferentially loaded the paretic leg during balance recovery^7^ or that elicited reactive stepping responses.^8^ Yet cortical activity after stroke has been assessed almost exclusively using the same perturbation magnitude across individuals, which imposes markedly different levels of balance challenge across individuals with different balance abilities.^9^ Assessing the scaling of cortical recruitment across graded levels of balance challenge^6,10–13^ could clarify how stroke constrains the neural strategies available for balance recovery, and what distinguishes individuals who achieve higher reactive balance performance.

The cortical N1 response is a negative electroencephalography (EEG) peak over midline sensorimotor areas approximately 150 ms after a balance disturbance, localized primarily to the supplementary motor area.^4,14–17^ The N1 is thought to reflect detection of an error between predicted and actual sensory information relevant to balance.^18–21^ N1 amplitudes increase and latencies shorten with perturbation magnitude,^4,6,10,11^ but scaling varies across individuals. Younger adults with worse balance show greater N1 scaling,^10^ and N1 scaling differs with long-term balance training.^13^ With aging, cortically mediated muscle activity is engaged at smaller perturbation magnitudes than in younger adults, suggesting that cortical resources may be recruited at lower levels of balance challenge.^5^ Relationships between N1 responses and balance behavior also differ between clinical and neurologically-intact populations.^7,12^ After stroke, we previously found smaller and delayed N1 responses than older adults at a small, standardized perturbation magnitude, with delayed responses associated with lower clinical balance function and slower reactive step initiation.^7,8^ However, whether these differences in cortical activity also influence how individuals scale cortical recruitment across different levels of balance challenge is unclear.

The Compensation Related Utilization of Neural Circuits Hypothesis (CRUNCH) describes greater recruitment of neural resources in older than younger adults at low task demand, and a lower ceiling on further recruitment as demand increases.^22^ CRUNCH extends to mobility, where older adults show greater recruitment and ceiling effects with increasing walking difficulty.^23,24^ After stroke, poorer mobility function is associated with greater prefrontal recruitment during typical walking and a lower recruitment ceiling during more demanding walking.^25,26^ In reactive balance, older adults with greater prefrontal-motor coherence during low-magnitude perturbations took reactive steps at smaller perturbation magnitudes, consistent with greater cortical engagement at low demand in individuals with lower reactive balance capacity.^9^ Whether cortical recruitment after stroke is similarly constrained as reactive balance challenge increases has not been tested.

Delivering perturbations at each individual’s reactive step threshold makes the predictions of CRUNCH testable during reactive balance recovery in older adults with and without stroke. Step threshold perturbations help to equalize balance challenge across people of differing balance ability, enabling within-subject comparisons across graded levels of challenge and between-subject comparisons at matched challenge.^13^ They additionally allow within-subject comparisons between stepping and feet-in-place reactions to the same perturbation stimulus.^27,28^ Larger N1 responses accompany reactive stepping,^4,28^ implicating differences in cortical error processing as a function of balance recovery strategy. However, the capacity to flexibly modulate cortical activity with changing motor demands is constrained after stroke,^29–31^ which may limit modulation of N1 responses between stepping and feet-in-place reactions.

The purpose of this study was to test predictions of CRUNCH during reactive balance recovery by comparing how older adults with and without stroke modulate cortical N1 responses across perturbation magnitudes and reactive balance outcomes. We predicted that N1 amplitudes would increase and latencies would shorten with perturbation magnitude in both groups,^4,6^ with greater N1 scaling in people with stroke given their lower clinical balance function.^10^ Consistent with CRUNCH, we further predicted that people with stroke with larger N1 responses at the smaller perturbation magnitude, reflecting over-recruitment of cortical resources, would show less N1 scaling at the larger magnitude, and that those with greater N1 scaling would reach higher step thresholds.^5,9,22^ Finally, we predicted that N1 responses would differ between feet-in-place and unintentional stepping responses to the same step threshold perturbation,^4,28^ and that this modulation would be reduced after stroke, given that the capacity to flexibly modulate cortical activity with changing motor demands is constrained after stroke.^29–31^

## Methods

### Participants and study procedures

The present study features a novel analysis of a reactive balance paradigm that involved an individualized perturbation series, with magnitudes scaled progressively based on each participant’s ability to recover balance without taking a step. We previously reported different analyses on a different subset of conditions from this same participant cohort.^7,9^ Eighteen individuals with chronic stroke (>6 months) and 17 age-similar controls completed a single-visit reactive balance assessment and clinical testing (i.e., Mini Balance Evaluation Systems Test (MiniBEST), Timed-Up-and-Go (TUG), and 10-meter walk test). Inclusion criteria included age >21 years, ability to walk at least 10 m without assistance, ability to stand unassisted for at least 3 minutes, and cognitive ability to provide informed consent. Participants were excluded if they had any neurological condition other than stroke, or pain affecting standing or walking. The clinical characteristics of this cohort are presented in Table 1, and the characteristics of the stroke lesion are described in Palmer et al. (2024).^7^ The Emory University Institutional Review Board approved the experimental protocol (IRB00083425), and all participants provided written informed consent.

**Table 1.**
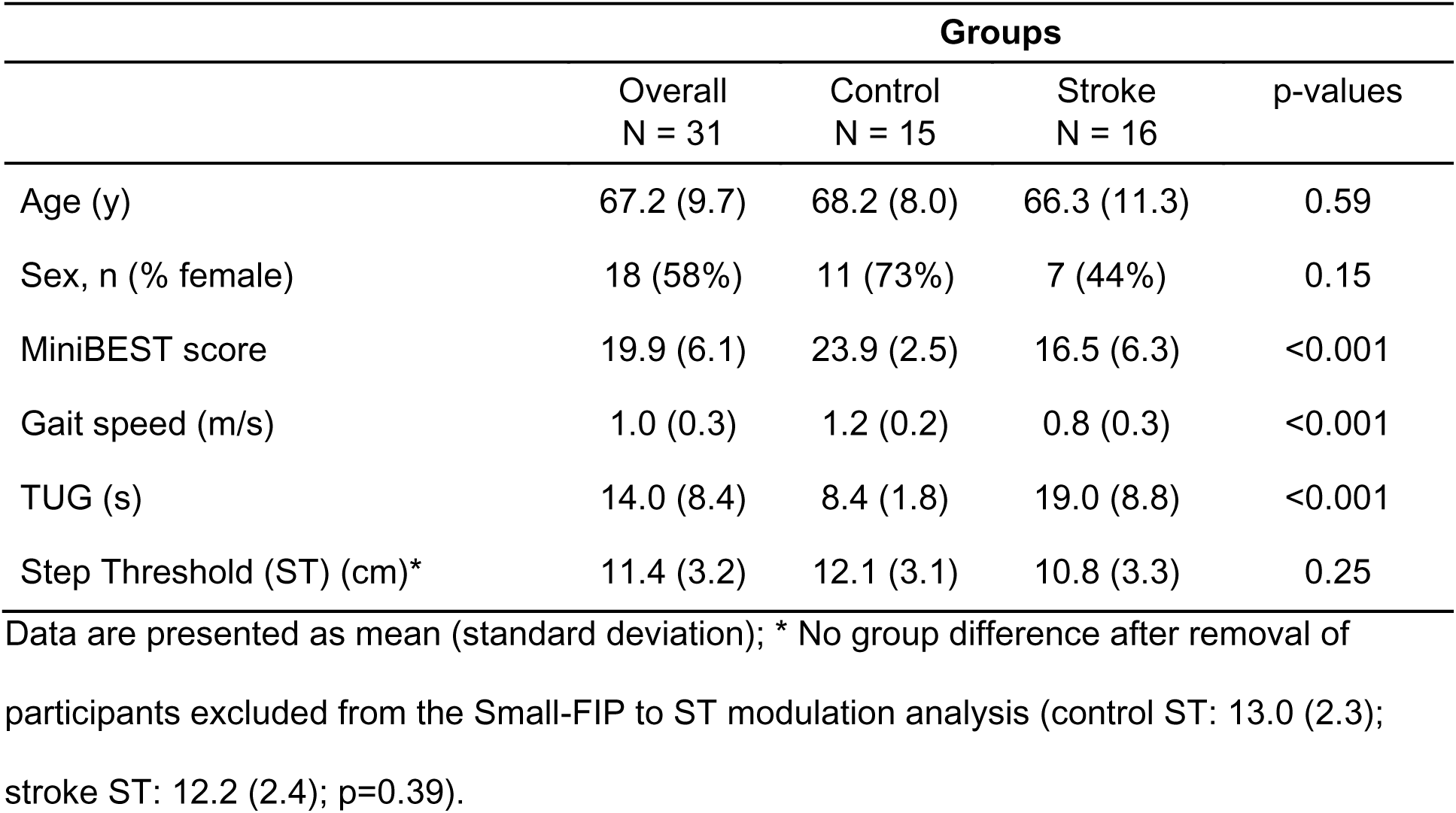
Demographics and clinical characteristics of the study cohort Data are presented as mean (standard deviation); * No group difference after removal of participants excluded from the Small-FIP to ST modulation analysis (control ST: 13.0 (2.3); stroke ST: 12.2 (2.4); p=0.39).

### Standing Balance Perturbations

Participants stood barefoot on a moving platform (Factory Automation Systems, Atlanta, GA) with arms folded across the chest and were instructed to recover balance with the feet in place if possible, stepping only when necessary to prevent a loss of balance. Participants first received a block of multidirectional perturbations at a small, standardized magnitude (7.5 cm, 16.0 cm/s, 0.12 g) that primarily elicited feet-in-place responses (Small-FIP).^7–9^ After a seated rest, participants received a second block of forward perturbations at their individual reactive step threshold (ST), defined as the magnitude eliciting an unintentional reactive step in approximately 50% of trials and identified by gradually escalating perturbation magnitudes.^9^ Occasional backward perturbations reduced anticipation of perturbation direction. Only forward perturbations at the small magnitude (Small-FIP) and at step threshold (ST-FIP and ST-step) were analyzed. Full protocol details are provided in the Supplemental Methods.

### EEG Data Acquisition and Analysis

EEG was recorded at 1000 Hz using a 64-channel active electrode system (actiCAP, actiCHamp, Brain Products GmbH, Gilching, Germany) and preprocessed in EEGLAB using the BeMoBIL pipeline,^32^ including line noise removal, bad channel detection and interpolation, and average referencing. Independent component analysis was performed using adaptive mixture independent component analysis (AMICA),^33^ and non-brain components were removed based on dipole fit residual variance, ICLabel classification, power spectral slope,^34^ TESA toolbox criteria,^35^ and visual inspection including power-power coupling.^36^ The final decomposition was back-projected onto data without the high-pass filter used for AMICA, to avoid distorting event-related potential waveforms.^37,38^ Data were then low-pass filtered (30 Hz), epoched, and baseline corrected (−150 to −50 ms before perturbation onset).^7,33^ Full preprocessing details are provided in the Supplemental Methods.

### Cortical N1 responses

N1 peak amplitude and latency were identified at Cz in each condition (Small-FIP, ST-FIP, ST-step) relative to each participant’s composite waveform across all forward perturbation trials (Supplemental Methods). N1 scaling (ΔN1 amplitude) and ΔN1 latency were quantified as the percent change from Small-FIP to ST-FIP, and the influence of behavioral outcome as the percent change from ST-FIP to ST-step at the same perturbation magnitude.

### Statistical analysis

Linear mixed models tested whether the effects of perturbation magnitude (Small-FIP, ST-FIP) and behavioral outcome (ST-FIP, ST-step) on N1 amplitude and latency differed between groups, with Bonferroni-corrected post hoc comparisons. Multiple linear regressions tested whether group moderated associations between N1 scaling and (1) Small-FIP N1 amplitude and (2) step threshold; because relationships between N1 responses and balance function differ between clinical and nonclinical populations,^7,12^ within-group standardized simple slopes were estimated a priori, and models refit without the interaction estimated associations across groups. Group comparisons, assumption checks, and additional details are provided in the Supplemental Methods; analyses used R (version 4.5.1; α = 0.05).

## Results

Sixteen people with stroke and 15 controls were included in the study analyses. Groups did not differ in age, sex, or step threshold (control: 12.1 ± 3.1 cm; stroke: 10.8 ± 3.3 cm; p = 0.25; **Table 1**, **Fig. 1A**, right panel), whereas the stroke group demonstrated lower miniBEST scores, slower self-selected gait speeds, and longer TUG times compared to controls (all p < 0.001; Table 1). One participant in the control group withdrew due to balance anxiety, one participant in the stroke group was unable to respond with FIP to forward perturbations, and two participants (one participant with stroke, one control) had technical issues with EEG data acquisition; these individuals were excluded from all reported analyses.

**Figure 1.**
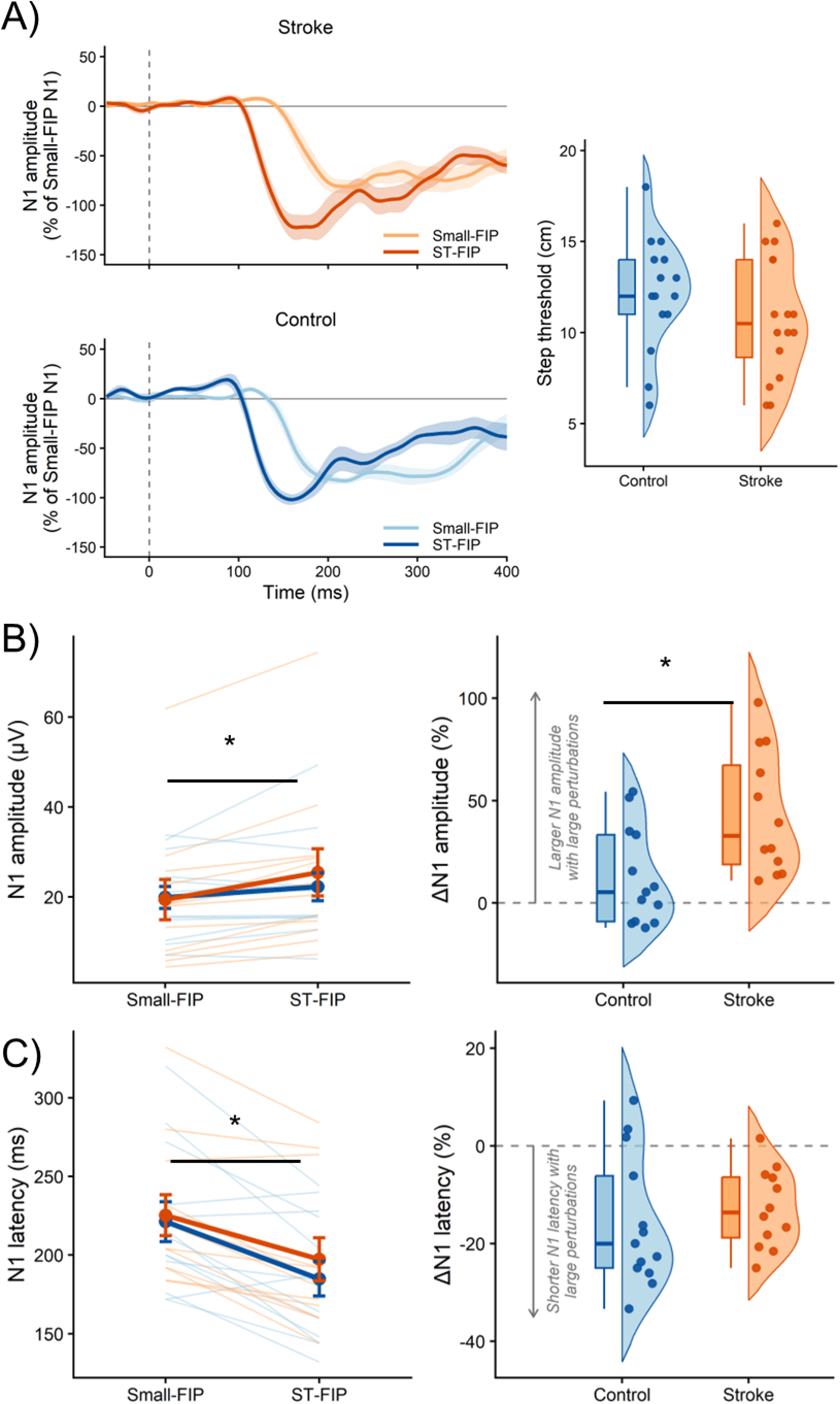
Perturbation magnitude effect on cortical N1 response scaling in stroke and control groups during feet-in-place (FIP) balance recovery responses. (A) Group-mean evoked cortical responses at Cz for the Small-FIP and step threshold (ST)-FIP conditions in the stroke (orange; n = 12) and control (blue; n = 13) groups (left panel). For each participant, waveforms from both conditions were normalized to that participant’s Small-FIP N1 peak amplitude before group averaging. The Small-FIP N1 peak therefore approximates 1, and the ST-FIP N1 peak reflects relative N1 scaling. Shaded ribbons indicate ±SEM; the dashed line denotes perturbation onset. Step threshold magnitude did not differ between groups across all participants (right panel; control, n = 15; stroke, n = 16; p = 0.25). (B) The N1 amplitude was larger during the ST-FIP compared to the Small-FIP conditions, regardless of group (p<0.001). People with stroke showed greater ΔN1 amplitude from the Small-FIP to ST-FIP conditions compared to controls (right panel) (p=0.009). (C) The N1 latency was shorter during the ST-FIP compared to Small-FIP conditions, regardless of group (p<0.001). There was no difference in ΔN1 latency between groups (p=0.51). * indicates statistical significance (p<0.05).

All 31 participants were included in comparisons of ST-FIP and ST-step trials, and 25 participants (13 control, 12 stroke) were included in comparisons across perturbation magnitudes. Six participants (n=2 control, n=4 stroke) were already at or above their step threshold in the smaller standardized perturbations; four of these participants had a step threshold that equaled the standardized small perturbation magnitude; for the other two, it was below it (7.0 vs. 7.5 cm). These six were therefore included in ST-FIP vs. ST-step analyses but not in comparisons across perturbation magnitudes. Step threshold also did not differ between groups within this subsample (control: 13.0 ± 2.3 cm; stroke: 12.2 ± 2.4 cm; p = 0.39). The number of trials retained per condition did not differ between groups (all p > 0.05). For the Small-FIP condition, the median number of trials was 24 (IQR = 0; range: 4–25) for the control group (n = 13) and 23 (IQR = 4; range: 17–24) for the stroke group (n = 12; W = 108, p = 0.062, r = 0.38). For the ST-FIP condition, the mean number of trials was 12.0 (SD = 7.2; range: 2–26) and 10.3 (SD = 4.5; range: 4–18) for the control (n = 15) and stroke (n = 16) groups, respectively (t(23.19) = 0.80, p = 0.43, d = 0.29). For the ST-step condition, the mean number of trials was 14.0 (SD = 7.6; range: 5–33) and 10.9 (SD = 6.2; range: 3–24) (t(27.18) = 1.22, p = 0.23, d = 0.44). All participants were retained in the main analyses regardless of trial count. As a sensitivity analysis, all data were reanalyzed after removing four participants with fewer than four trials in the ST-FIP (n = 2) or ST-step (n = 2) condition; these analyses yielded results similar to those reported below.

### Cortical N1 amplitude scales more with perturbation magnitude in people with stroke than controls

During feet-in-place balance recovery, peak N1 responses did not differ between groups but became larger and earlier at step threshold than at the small perturbation magnitude in both groups. There was no difference in N1 amplitude (F(1, 26.98) = 0.32, p = 0.57) or latency (F(1, 26.90) = 1.20, p = 0.28) between groups. The interaction effect between perturbation magnitude and group was not significant for N1 amplitude (F(1, 23.14) = 3.58, p = 0.071) (Fig. 1B, left panel) or N1 latency (F(1, 23.43) = 0.92, p = 0.35) (Fig. 1C, left panel). Larger perturbations elicited greater N1 amplitudes (Small-FIP = 18.5 ± 2.61 µV, ST-FIP = 22.6 ± 2.59 µV) (F(1, 23.14) = 20.53, p < 0.001) (**Fig. 1B**, left panel) and shorter N1 latencies (Small-FIP = 227 ± 8.6 ms, ST-FIP = 196 ± 8.4 ms) (F(1, 23.43) = 35.93, p < 0.001) (**Fig. 1C**, left panel).

People with stroke showed a greater relative increase in N1 amplitude from the Small-FIP to the ST-FIP condition than controls (ΔN1 amplitude: stroke mean = 43.5%, SD = 30.0%; control mean = 12.5%, SD = 23.6%; t(20.91) = −2.86, p = 0.009, d = −1.16; **Fig. 1B**, right panel). This group difference was also present for the absolute change in N1 amplitude (stroke median = 5.22 µV, IQR = 4.94; control median = 0.80 µV, IQR = 5.63; W = 36, p = 0.024, r = 0.54), and every participant with stroke increased N1 amplitude from the Small-FIP to the ST-FIP condition (12 of 12) compared with 8 of 13 controls (p =0.039, Fisher’s exact test). In contrast, groups did not differ in ΔN1 latency (t_19.86_ = −0.67, Cohen’s d = −0.26, p = 0.51) (**Fig. 1C**, right panel), nor in the raw change in N1 latency (stroke = −28.0 ± 18.0 ms, control = −36.3 ± 32.1 ms; t(19.14) = −0.81, Cohen’s d = −0.32, p = 0.43).

### Greater cortical N1 scaling is associated with higher step thresholds across groups

N1 amplitude scaling was greater in participants with stroke who had smaller N1 amplitudes in the Small-FIP condition. Within the stroke group, lower Small-FIP N1 amplitudes were associated with greater N1 amplitude scaling (β = −0.43, 95% CI [−0.83, −0.02], p = 0.041, **Fig. 2A**), whereas no association was present in controls (β = −0.07, 95% CI [−0.76, 0.62], p = 0.83). However, these associations did not differ significantly between groups (group × Small-FIP N1 amplitude interaction: β = −0.36, 95% CI [−1.16, 0.45], p = 0.37). With the interaction term removed, Small-FIP N1 amplitude was not associated with N1 amplitude scaling across both groups (β = −0.33, 95% CI [−0.68, 0.01], p = 0.060), whereas participants with stroke showed greater N1 amplitude scaling than controls (β = 1.00, 95% CI [0.31, 1.68], p = 0.006; adjusted R² = 0.32, F(2,22) = 6.67, p = 0.005).

**Figure 2.**
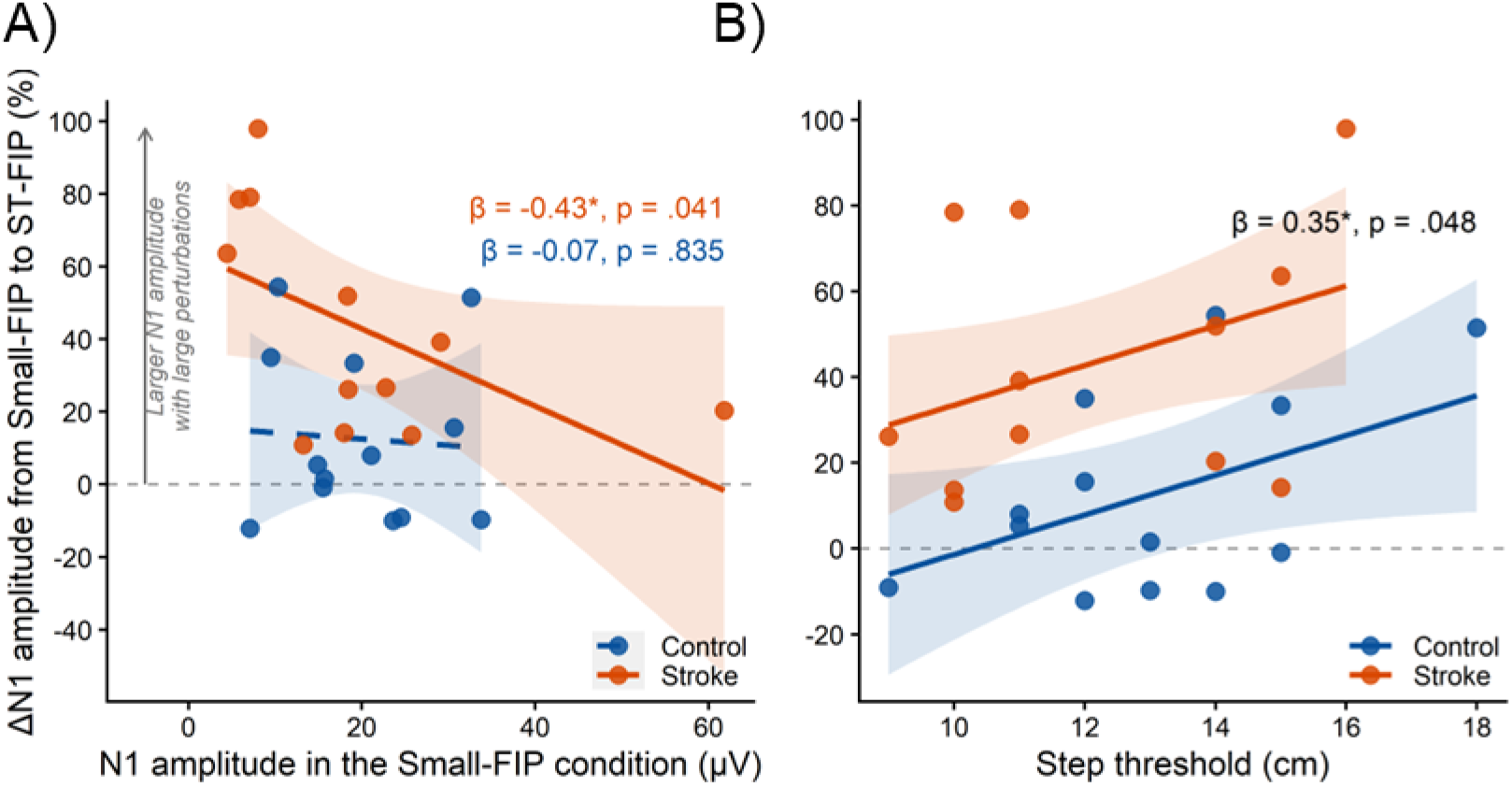
Cortical N1 modulation across stroke and control groups with respect to N1 responses at small perturbation magnitudes and step threshold balance performance. **(A)** The relationship between scaling of N1 amplitude (ΔN1 amplitude from the Small-FIP to the ST-FIP conditions) and N1 amplitude in the Small-FIP condition did not differ between people with stroke and controls (t=−0.92, p=0.37). People after stroke who elicited smaller N1s during Small-FIP perturbations showed greater scaling of N1 amplitude with larger ST perturbations (β=−0.43, 95%CI, [−0.83, −0.02], p=0.041), while controls showed no relationship (β=−0.07, 95%CI, [−0.76, 0.62], p=0.83). (B) People with higher step thresholds (indicating greater reactive balance performance) showed greater scaling of N1 amplitude with larger ST perturbations, regardless of group (β=0.35, 95%CI, [0.003, 0.71], p=0.048). Lines in (A) show within-group fits; lines in (B) show the group-adjusted fit (common slope) for each group. Shaded bands are 95% confidence intervals, and dashed lines indicate a non-significant slope. * indicates p<0.05.

Across both groups, participants with higher step thresholds showed greater N1 amplitude scaling. The association between step threshold and N1 scaling did not differ between groups (group × step threshold interaction: β = −0.12, 95% CI [−0.84, 0.60], p = 0.73) and was not significant within either group alone (stroke: β = 0.30, 95% CI [−0.21, 0.80], p = 0.23; control: β = 0.42, 95% CI [−0.10, 0.93], p = 0.11). With the interaction term removed, a higher step threshold was associated with greater N1 amplitude scaling across both groups (β = 0.35, 95% CI [0.003, 0.71], p = 0.048; adjusted R² = 0.33, F(2,22) = 6.95, p = 0.005; **Fig. 2B**) and participants with stroke showed greater N1 amplitude scaling than controls (β = 1.14, 95% CI [0.45, 1.83], p = 0.002; adjusted R² = 0.33, F(2, 22) = 6.95, p = 0.005). In contrast, N1 amplitude scaling was not associated with age, Mini-BESTest score, self-selected gait speed, or TUG time after accounting for group (all p > 0.6), and none of these associations differed between groups (all group × measure interactions p ≥ 0.19).

### Cortical N1 timing but not amplitude differs with balance recovery outcome in controls but not in people with stroke

In controls but not in people with stroke, trials recovered with a reactive step (unsuccessful task performance) at the step threshold perturbation magnitude elicited longer N1 latencies than trials recovered with the feet in place (successful task performance), whereas N1 amplitude did not differ between these behavioral outcomes. There was no significant interaction between behavioral outcome (ST-FIP, ST-step) and group for N1 amplitude (F(1, 29) = 3.58, p = 0.068; **Fig. 3B**, left panel), and ΔN1 amplitude from the ST-FIP to the ST-step conditions did not differ between groups (t(28.56) = 1.83, p = 0.078, d = 0.66; Fig. 3B, right panel). For N1 latency, there was a significant interaction between group and behavioral outcome (F(1, 29) = 5.65, p = 0.024; **Fig. 3C**, left panel). N1 latencies were longer during ST-step compared to ST-FIP trials in the control group (mean difference ± SE = 11.2 ± 2.63 ms, p < 0.001) but not the stroke group (mean difference = 2.50 ± 2.55 ms, p = 0.67). Consistent with this, the control group showed a greater ΔN1 latency from ST-FIP to ST-step than the stroke group (control: median = 4.3%, IQR = 6.47%; stroke: median = 0%, IQR = 2.20%; W = 193, p = 0.004, r = 0.61; Fig. 3C, right panel).

**Figure 3.**
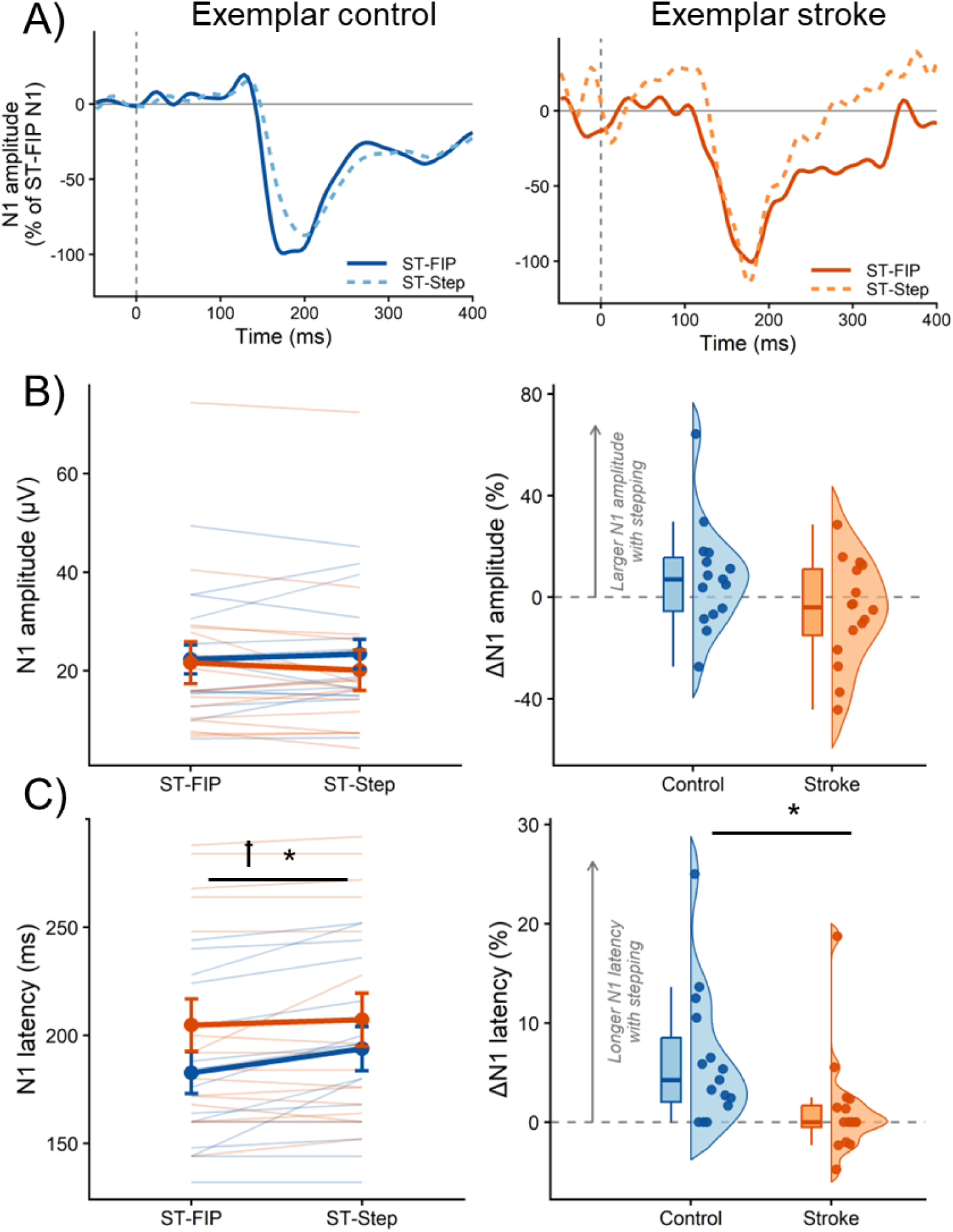
Behavioral effect of balance recovery responses on cortical N1 response scaling in stroke and control groups during balance recovery at large step threshold (ST) perturbations. (A) Mean evoked cortical N1 responses during ST-FIP and ST-step behaviors for an exemplar older adult with stroke (orange) and without stroke (blue). The dashed line denotes the onset of the perturbation. (B) There was no interaction effect of group-by-behavior on N1 amplitude (p=0.07), nor a significant group-level difference in ΔN1 amplitude (p=0.08) (right panel). (C) The control group showed a longer N1 latency during the ST-step (unsuccessful) compared to the ST-FIP (successful) task behavior (p<0.001), while the stroke group showed no difference (p=0.67) (left panel). The control group showed greater ΔN1 latency between the ST-FIP and ST-step behaviors compared to the stroke group (p=0.004). ꝉ indicates a significant interaction effect (p=0.024), while * represents simple effects differences in the control group between balance behaviors of step and FIP responses (p<0.001).

## Discussion

People with stroke may compensate for impaired automatic balance control by engaging greater cortical resources as balance demands increase, and this engagement appears to be adaptive rather than solely a marker of neurologic impairment. Despite lower clinical balance and mobility scores, people with stroke achieved reactive step thresholds comparable to those of older adults without stroke while showing a greater relative increase in cortical N1 amplitude from the small to the step threshold perturbation magnitude. Across both groups, greater cortical scaling was associated with higher step thresholds, suggesting that the capacity to increase cortical engagement with perturbation magnitude may support reactive balance performance. In people with stroke, a higher step threshold may itself reflect behavioral compensation, favoring feet-in-place recovery when reactive stepping with the paretic leg is delayed or unreliable. At step threshold, the timing of cortical responses differed with behavioral outcome in older adults without stroke, with later N1 peaks on trials recovered with a reactive step than on trials recovered with the feet in place, whereas cortical response timing in people with stroke was similar regardless of outcome. Together, these findings suggest that comparable reactive balance performance after stroke may be achieved through different patterns of cortical engagement, which become evident when perturbation magnitudes are scaled to individual balance capacity.

Group differences in the cortical N1 response may diminish as balance challenge approaches the limits of reactive balance performance, whereas increasing balance challenge may drive greater within-person modulation of cortical responses in people with stroke. The cortical N1 response is thought to reflect cortical assessment of balance error, increasing in amplitude with perturbation magnitude^4,6^ and postural threat,^39^ and in people with lower balance ability.^10^ In our previous report in this cohort, people with stroke showed smaller and later N1 responses than controls during backward support-surface translations that displaced the center of mass forward, toward the toes.^7^ Forward support-surface translations, which displace the center of mass backward toward the heels, are more biomechanically challenging.^40^ During these more challenging forward translations in the present study, N1 amplitude and latency did not differ between groups. Within each person, however, N1 amplitude typically increased from the small to the step threshold perturbation magnitude, and this increase was greater in people with stroke (43.5% vs. 12.5%). Every participant with stroke increased N1 amplitude, compared with 8 of 13 controls. Because step thresholds did not differ between groups, greater N1 scaling after stroke cannot be attributed to larger perturbations. Given that N1 amplitude is thought to reflect cortical assessment of balance error,^6,10^ greater scaling may indicate that balance at step threshold is interpreted as a larger balance error by people with stroke. Together, these findings suggest that balance challenge is a key driver of within-person modulation of balance error responses after stroke.

Greater cortical scaling after stroke may reflect greater use of an adaptive reactive balance strategy shared with older adults without stroke, rather than solely a marker of impairment. Across both groups, people with higher step thresholds showed greater N1 scaling (Fig. 2B), similar to the relationship observed in skilled dancers.^13^ Because step threshold determines the larger perturbation magnitude, this relationship may partly reflect the size of the perturbation increase. However, greater scaling in people with stroke persisted after accounting for step threshold (Fig. 2B), indicating that the group difference does not arise from differences in perturbation magnitude. The comparable step thresholds were unexpected given the lower clinical balance and mobility scores in the stroke group (Table 1), and suggest that step threshold may index compensation as well as biomechanical reactive balance capacity after stroke. Delayed reactive step initiation after stroke^8^ could make stepping a slower and less reliable option, favoring feet-in-place recovery after stroke. In contrast, older adults without stroke may need to inhibit a prepotent stepping response to recover with the feet in place, consistent with greater midfrontal theta activity during instructed feet-in-place balance recovery in young adults.^27^ A feet-in-place response may therefore be favored after stroke even when it is more posturally demanding, and maintaining it may require cortical engagement. These findings are only partly consistent with the CRUNCH model, which describes over-recruitment of neural resources at low task demand and a ceiling on further recruitment as demand increases.^22–24^ Consistent with CRUNCH, the capacity to increase cortical engagement with task demand was associated with better performance. However, people with stroke did not show larger N1 responses than controls at the small perturbation magnitude, and they increased N1 amplitude more, not less, than controls; this result contrasts with group-level over-recruitment and restricted range observed in older adults during uneven terrain walking.^24^ Within the stroke group, people with smaller N1 responses at the small perturbation magnitude showed greater scaling (Fig. 2A), which may reflect greater reserve for cortical recruitment. However, this association did not differ significantly from that in controls and should be interpreted cautiously. Rather than reflecting a limit on cortical resources, greater cortical scaling after stroke under challenging reactive balance conditions may reflect the capacity to engage adaptive neural strategies in the presence of motor impairment. Longitudinal studies are needed to determine whether this capacity emerges during stroke recovery to support reactive balance.

Greater cortical scaling may reflect heightened sensitivity of cortical balance error assessment, which may support reactive balance at both ends of a U-shaped relationship between cortical engagement and balance skill. Modern dancers show larger N1 responses than nondancers, interpreted as heightened sensitivity of error assessment supporting precise postural control rather than compensation for poor balance.^13,41,42^ Like dancers, people in the present study with higher step thresholds showed greater N1 scaling,^13^ suggesting that sensitivity to increasing balance error supports reactive balance across levels of skill. In people with stroke, heightened sensitivity may instead develop as an adaptation to chronically high balance demands imposed by hemiparesis. These findings have important clinical implications for perturbation-based balance training, which has shown effectiveness in people after stroke.^43–45^ Whether training acts by increasing scaling capacity or by reducing cortical engagement at low challenge remains unknown, and measuring cortical responses before and after training could distinguish these possibilities.

Reactive balance strategy may depend on the precise timing of cortical error assessment, a coupling between cortical timing and behavior that appears dissociated after stroke. In controls, but not in people with stroke, N1 peaks were later on trials recovered with a reactive step than on trials recovered with the feet in place, whereas N1 amplitude did not differ between outcomes. The N1 is thought to reflect cortical assessment of balance error^6,19,20^ and peaked before reactive step initiation in this cohort.^8^ Later balance error assessment on a given trial may therefore limit the ability to inhibit a prepotent step,^27^ rather than reflecting a consequence of stepping. In people with stroke, N1 timing did not vary with behavioral outcome. However, later N1 responses have been associated with lower clinical balance and mobility function^7^ and slower reactive step initiation^8^ after stroke. The timing of cortical error assessment may thus influence which recovery strategy is selected on a given trial in older adults, and in contrast may influence how quickly any reactive response is initiated after stroke. Whether this balance N1-behavioral coupling can be shaped through balance training, as differences between individuals with and without long-term balance training might suggest,^13^ is an important next step for future research with direct implications for post-stroke rehabilitation.

### Limitations

Our sample and design constrain the generalizability of these findings. The cross-sectional, case-control design precludes causal inference, and the small sample warrants confirmation in larger cohorts with more trials per condition. Participants who reached step threshold at or below the small standardized magnitude could not be included in the scaling analyses, so these analyses represent a relatively higher-functioning subsample of the stroke cohort. Stroke lesion location and volume could not be modeled robustly in a sample of this size, yet may meaningfully influence cortical responses. Finally, comparison with prior work is complicated by differing experimental paradigms, including perturbation direction and predictability and whether stepping is cued or spontaneous, and by evidence that relationships between cortical responses and balance behavior differ between clinical and neurologically-intact populations.^12^

Several methodological factors may have influenced N1 measurement across conditions. Perturbation conditions were delivered in a fixed order, with the small perturbation block preceding the step threshold block, and we did not quantify N1 habituation over repeated perturbations.^6,15^ Because habituation would attenuate responses more in the later step threshold block, it would reduce rather than inflate the observed N1 increase with perturbation magnitude. Tolerance for repeated high-magnitude perturbations also limited the number of trials at step threshold. Identifying condition-specific N1 peaks relative to a within-participant composite waveform likely improved the consistency of latency identification, but does not eliminate the influence of trial number on the reliability of N1 latency quantification.^33^ Finally, our channel-based approach to N1 quantification has excellent reliability^33^ but cannot delineate neural source contributions to the N1, an important direction for identifying neural targets for rehabilitation.^16,17^

## Conclusions

Consistent with our hypothesis, people with stroke showed greater cortical engagement as perturbation magnitude increased, and greater engagement was associated with higher reactive step thresholds across groups, suggesting an adaptive strategy rather than solely a marker of impairment. In contrast, the timing of cortical error assessment was less coupled to balance recovery strategy after stroke. Comparable reactive balance after stroke may therefore be achieved through different patterns of cortical engagement, which become apparent when perturbations are individualized to balance capacity beyond what clinical balance assessment captures. Neurorehabilitation interventions that expand the scaling of cortical resources engaged across levels of balance challenge, or that restore precision and flexibility of balance error response modulation, may represent complementary targets for improving reactive balance capacity after stroke.

## Data Availability

Data produced in the present study are available upon reasonable request to the authors.

## Acknowledgments

We would like to acknowledge Michelle Goto, Pam Inglett, and Annie Goldman for their assistance with data collection for this project. We thank members of the Brains in Motion Laboratory at the University of Minnesota for discussions that contributed to the idea of extending prior analyses in dancers to examine relationships between N1 scaling and step threshold in people with stroke.

## Author Contributions

Jacqueline A. Palmer: Conceptualization; Data curation; Formal analysis; Funding acquisition; Investigation; Methodology; Project administration; Resources; Software; Visualization; Writing – original draft; Writing – review & editing. Lena H. Ting: Conceptualization; Funding acquisition; Writing – review & editing. Michael R. Borich: Conceptualization; Funding acquisition; Writing – review & editing. Aiden M. Payne: Data curation; Formal analysis; Investigation; Methodology; Software; Writing – review & editing. Jasmine L. Mirdamadi: Formal analysis; Software; Writing – review & editing.

## Statements and Declarations

### Ethical considerations

The Emory University Institutional Review Board approved the experimental protocol (IRB00083425).

## Consent to participate

All participants provided written informed consent prior to participation.

## Consent for publication

Not applicable.

## Data availability

The data that support the findings of this study are available from the corresponding author upon reasonable request.

## Declaration of generative AI use

Generative AI (Claude, Anthropic) was used to assist with editing and condensing manuscript text, including the presentation of statistical methods, to provide pre-submission review feedback, and to check consistency between in-text citations and the reference list. The authors reviewed, revised, and verified all AI-assisted text. Study design, data analysis, interpretation, and conclusions were developed by the authors, who take full responsibility for the content of this manuscript.

## References

1. Harris J, Eng J, Marigold D, Tokuno C, Louis C. Relationship of balance and mobility to fall incidence in people with chronic stroke. Phys Ther. 2005;85(2):150–158.

2. Mansfield A, Wong JS, McIlroy WE, et al. Do measures of reactive balance control predict falls in people with stroke returning to the community? Physiotherapy. 2015;101(4):373–380. doi:10.1016/j.physio.2015.01.009

3. Horak FB, Jacobs JV. Cortical control of postural responses. J Neural Transm. 2007;114(10):1339–1348. doi:10.1007/s00702-007-0657-0.Cortical

4. Solis-Escalante T, Stokkermans M, Cohen MX, Weerdesteyn V. Cortical responses to whole-body balance perturbations index perturbation magnitude and predict reactive stepping behavior. Eur J Neurosci. Published online September 15, 2020. doi:10.1111/ejn.14972

5. Boebinger SE, Payne AM, Xiao J, et al. Cortically Mediated Muscle Responses to Balance Perturbations Increase with Perturbation Magnitude in Older Adults with and without Parkinson’s Disease. eNeuro. 2026;13(4):ENEURO.0423-25.2026. doi:10.1523/ENEURO.0423-25.2026

6. Payne AM, Hajcak G, Ting LH. Dissociation of muscle and cortical response scaling to balance perturbation acceleration. J Neurophysiol. 2019;121(3):867–880. doi:10.1152/jn.00237.2018

7. Palmer JA, Payne AM, Mirdamadi JL, Ting LH, Borich MR. Delayed Cortical Responses During Reactive Balance After Stroke Associated With Slower Kinetics and Clinical Balance Dysfunction. Neurorehabil Neural Repair. Published online September 27, 2024:15459683241282786. doi:10.1177/15459683241282786

8. Tsai SY, Payne AM, Mirdamadi JL, Ting LH, Borich MR, Palmer JA. Slower reactive stepping kinematics are associated with lower clinical balance function and delayed cortical evoked responses under challenging balance conditions after stroke. medRxiv. Preprint posted online December 19, 2025:2025.12.18.25342582. doi:10.64898/2025.12.18.25342582

9. Palmer JA, Payne AM, Ting LH, Borich MR. Cortical Engagement Metrics During Reactive Balance Are Associated With Distinct Aspects of Balance Behavior in Older Adults. Front Aging Neurosci. 2021;13:410. doi:10.3389/fnagi.2021.684743

10. Payne AM, Ting LH. Worse balance is associated with larger perturbation-evoked cortical responses in healthy young adults. Gait Posture. 2020;80:324–330.

11. Payne AM, Palmer JA, McKay JL, Ting LH. Lower Cognitive Set Shifting Ability Is Associated With Stiffer Balance Recovery Behavior and Larger Perturbation-Evoked Cortical Responses in Older Adults. Front Aging Neurosci. 2021;13:742243. doi:10.3389/fnagi.2021.742243

12. Payne AM, McKay JL, Ting LH. The cortical N1 response to balance perturbation is associated with balance and cognitive function in different ways between older adults with and without Parkinson’s disease. Cereb Cortex Commun. 2022;3(3):tgac030. doi:10.1093/texcom/tgac030

13. Kerr KG, Boebinger SE, Mirdamadi JL, Protzak J, Borich MR, Ting LH. Cortical response to balance perturbation is more sensitive in modern dancers than nondancers during biomechanically similar balance recovery. J Neurophysiol. Published online February 17, 2026. doi:10.1152/jn.00606.2025

14. Marlin A, Mochizuki G, Staines WR, McIlroy WE. Localizing evoked cortical activity associated with balance reactions: does the anterior cingulate play a role? J Neurophysiol. 2014;111(12):2634–2643. doi:10.1152/jn.00511.2013

15. Mierau A, Hülsdünker T, Strüder HK. Changes in cortical activity associated with adaptive behavior during repeated balance perturbation of unpredictable timing. Front Behav Neurosci. 2015;9(October):1–12. doi:10.3389/fnbeh.2015.00272

16. Mirdamadi JL, Ting LH, Borich MR. Distinct Cortical Correlates of Perception and Motor Function in Balance Control. J Neurosci Off J Soc Neurosci. 2024;44(15):e1520232024. doi:10.1523/JNEUROSCI.1520-23.2024

17. Protzak J, Mirdamadi JL, Borich MR, Ting LH. A primary central source determines perturbation-evoked N1 amplitudes in younger adults. J Neurophysiol. 2026;135(5):1315–1323. doi:10.1152/jn.00002.2026

18. Adkin AL, Quant S, Maki BE, McIlroy WE. Cortical responses associated with predictable and unpredictable compensatory balance reactions. Exp Brain Res. 2006;172(1):85–93. doi:10.1007/s00221-005-0310-9

19. Payne AM, Ting LH, Hajcak G. Do sensorimotor perturbations to standing balance elicit an error-related negativity? Psychophysiology. 2019;56(7):1–12. doi:10.1111/psyp.13359

20. Payne AM, Ting LH, Hajcak G. The balance N1 and the ERN correlate in amplitude across individuals in small samples of younger and older adults. Exp Brain Res. 2023;241(10):2419–2431. doi:10.1007/s00221-023-06692-9

21. Jansen DNR, Mensink L, Leeuwis M, Forbes PA. Cortical responses to balance perturbations persist without active postural control. J Physiol. 2026;604(10):3914–3933. doi:10.1113/JP290280

22. Reuter-Lorenz PA, Cappell KA. Neurocognitive Aging and the Compensation Hypothesis. Curr Dir Psychol Sci. 2008;17(3):177–182. doi:10.1111/j.1467-8721.2008.00570.x

23. Clark DJ, Manini TM, Ferris DP, et al. Multimodal Imaging of Brain Activity to Investigate Walking and Mobility Decline in Older Adults (Mind in Motion Study): Hypothesis, Theory, and Methods. Front Aging Neurosci. 2019;11:358. doi:10.3389/fnagi.2019.00358

24. Pliner EM, Liu C, Salminen JS, et al. Compensation Related Utilization of Neural Circuits Hypothesis (CRUNCH) of Electrocortical Data during Walking on Uneven Terrain. PLOS Aging Health. 2026;1(1):e0000017. doi:10.1371/journal.page.0000017

25. Hawkins KA, Fox EJ, Daly JJ, et al. Prefrontal over-activation during walking in people with mobility deficits: Interpretation and functional implications. Hum Mov Sci. 2018;59:46–55. doi:10.1016/j.humov.2018.03.010

26. Chatterjee SA, Fox EJ, Daly JJ, et al. Interpreting Prefrontal Recruitment During Walking After Stroke: Influence of Individual Differences in Mobility and Cognitive Function. Front Hum Neurosci. 2019;13:194. doi:10.3389/fnhum.2019.00194

27. Teodoro Solis-Escalante; Weerdesteyn V. Cortical dynamics during preparation and execution of reactive balanceresponses with distinct postural demands. NeuroImage. Published online 2019:1–15.

28. Payne AM, Ting LH. Balance perturbation-evoked cortical N1 responses are larger when stepping and not influenced by motor planning. J Neurophysiol. 2020;124(6):1875–1884. doi:10.1152/jn.00341.2020

29. Palmer JA, Kesar TM, Wolf SL, Borich MR. Motor Cortical Network Flexibility is Associated With Biomechanical Walking Impairment in Chronic Stroke. Neurorehabil Neural Repair. Published online September 27, 2021:15459683211046272. doi:10.1177/15459683211046272

30. Palmer JA, Wheaton LA, Gray WA, Silva MAS da, Wolf SL, Borich MR. Role of Interhemispheric Cortical Interactions in Poststroke Motor Function: *Neurorehabil Neural Repair*. Published online July 22, 2019. doi:10.1177/1545968319862552

31. Rossiter HE, Boudrias M hélène, Ward NS. Do movement-related beta oscillations change after stroke ? J Neurophysiol. 2014;112(9):2053–2058. doi:10.1152/jn.00345.2014

32. Klug M, Jeung S, Wunderlich A, et al. The BeMoBIL Pipeline for automated analyses of multimodal mobile brain and body imaging data. *bioRxiv*. Preprint posted online October 6, 2022:2022.09.29.510051. doi:10.1101/2022.09.29.510051

33. Mirdamadi JL, Poorman A, Munter G, et al. Excellent test-retest reliability of perturbation-evoked cortical responses supports feasibility of the balance N1 as a clinical biomarker. J Neurophysiol. 2025;133(3):987–1001. doi:10.1152/jn.00583.2024

34. Fitzgibbon SP, DeLosAngeles D, Lewis TW, et al. Automatic determination of EMG-contaminated components and validation of independent component analysis using EEG during pharmacologic paralysis. Clin Neurophysiol Off J Int Fed Clin Neurophysiol. 2016;127(3):1781–1793. doi:10.1016/j.clinph.2015.12.009

35. Rogasch NC, Sullivan C, Thomson RH, et al. Analysing concurrent transcranial magnetic stimulation and electroencephalographic data: A review and introduction to the open-source TESA software. NeuroImage. 2017;147:934–951. doi:10.1016/j.neuroimage.2016.10.031

36. Thammasan N, Miyakoshi M. Cross-Frequency Power-Power Coupling Analysis: A Useful Cross-Frequency Measure to Classify ICA-Decomposed EEG. Sensors. 2020;20(24):7040. doi:10.3390/s20247040

37. de Cheveigné A, Nelken I. Filters: When, Why, and How (Not) to Use Them. Neuron. 2019;102(2):280–293. doi:10.1016/j.neuron.2019.02.039

38. Tanner D, Morgan-Short K, Luck SJ. How inappropriate high-pass filters can produce artifactual effects and incorrect conclusions in ERP studies of language and cognition. Psychophysiology. 2015;52(8):997–1009. doi:10.1111/psyp.12437

39. Adkin AL, Campbell AD, Chua R, Carpenter MG. The influence of postural threat on the cortical response to unpredictable and predictable postural perturbations. Neurosci Lett. 2008;435(2):120–125. doi:10.1016/j.neulet.2008.02.018

40. Pai YC, Patton J. Center of mass velocity-position predictions for balance control. J Biomech. 1997;30(4):347–354. doi:10.1016/s0021-9290(96)00165-0

41. Guadagnoli MA, Lee TD. Challenge point: a framework for conceptualizing the effects of various practice conditions in motor learning. J Mot Behav. 2004;36(2):212–224. doi:10.3200/JMBR.36.2.212-224

42. Woollacott M, Shumway-Cook A. Attention and the control of posture and gait: a review of an emerging area of research. Gait Posture. 2002;16(1):1–14. doi:10.1016/S0966-6362(01)00156-4

43. Bhatt T, Dusane S, Patel P. Does severity of motor impairment affect reactive adaptation and fall-risk in chronic stroke survivors? J NeuroEngineering Rehabil. 2019;16(1):1–13. doi:10.1186/s12984-019-0510-3

44. Bhatt T, Dusane S, Gangwani R, Wang S, Kannan L. Motor adaptation and immediate retention to overground gait-slip perturbation training in people with chronic stroke: an experimental trial with a comparison group. Front Sports Act Living. 2023;5:1195773. doi:10.3389/fspor.2023.1195773

45. Patel PJ, Bhatt T. Does aging with a cortical lesion increase fall-risk: Examining effect of age versus stroke on intensity modulation of reactive balance responses from slip-like perturbations. Neuroscience. 2016;333:252–263. doi:10.1016/j.neuroscience.2016.06.044

