## Supplemental Methods for "Greater scaling and less flexible timing of cortical responses during challenging reactive balance after stroke"

### Standing Balance Perturbations

Participants stood barefoot on a moving platform level with the floor with feet approximately shoulder-width apart and arms folded across the chest. We delivered support-surface translational perturbations using the moving platform (Factory Automation Systems, Atlanta, GA). Participants were instructed to stand in their self-selected posture, to recover their balance using a feet-in-place strategy if possible, and to take a reactive step only when necessary to prevent a loss of balance. Participants first underwent a block of multidirectional perturbations at a small, standardized perturbation magnitude (7.5 cm, 16.0 cm/s, 0.12 g) that primarily elicited feet-in-place responses (Small-FIP), with a time interval between perturbations ranging between 15-60 seconds.^7–9^ After a seated rest break (5-7 minutes), participants underwent a second block of anteriorly directed perturbations that were scaled to an individualized magnitude at the reactive step threshold (ST) for each participant. First, the step threshold was defined as the perturbation magnitude that elicited an unintentional reactive stepping response in approximately 50% of the trials, identified by gradually escalating perturbation magnitudes.^9^ Posteriorly directed perturbations were delivered once approximately every 10 trials to minimize participant neuromuscular and biomechanical anticipation of the perturbation direction. Only trials delivered in the forward direction at small perturbations (Small-FIP) and step threshold (ST-FIP and ST-step) were included in the current analysis. Participants rested in a seated position every eight minutes or more frequently upon request. Following any trial in which a reactive step occurred, the experimenter confirmed that foot placement had returned to the baseline position before initiating the subsequent trial.

### EEG Data Acquisition and Analysis

Cortical activity during standing balance perturbations was continuously recorded (sampling rate 1000Hz) using EEG, with a 64-channel active electrode cap (actiCAP, actiCHamp amplifier, Brain Products, GmbH, Gilching, Germany). EEG signals were digitized using the actiCHamp's integrated 24-bit A/D converter and an online 20 kHz antialiasing low-pass filter (Brain Products, GmbH).

Raw EEG data were first down-sampled to 250Hz and preprocessed in EEGLAB using the BeMoBIL pipeline for mobile brain/body imaging data.^32^ The BeMoBIL preprocessing pipeline consisted of line noise removal using the Zapline-plus plugin and bad channel detection and removal using the clean_rawdata plugin. Since the clean_rawdata plugin uses random data sampling, the channel rejection procedure was repeated for twenty iterations. For each iteration, channels that correlated with nearby channels <0.7 were flagged as bad channels. A channel was removed if it was flagged as “bad” in more than 7 out of 20 iterations. Bad channels were subsequently interpolated, then the data were average referenced including interpolated channels. Preprocessed data were then high-pass filtered with a cut-off frequency of 1.75 Hz to optimize the adaptive mixture component analysis algorithm (AMICA) analyses. Then AMICA was performed on trial-epoched data (-2 to 2s).^33^ ICs were removed based on the following criteria: 1) any ICs with high dipole fit residual variance (>25%), 2) ICs classified by ICLabel as eye activity, cardiac activity, or line or channel noise with the highest probability, 3) ICs in which the linear slope of the power spectrum between 2 and 40 Hz was greater than 0.33, as commonly seen in muscle artifacts,^34^ and 4) any additional ICs labeled as ‘bad channels’ using the TESA toolbox.^35^ ICs identified as non-brain sources were verified by visual inspection before removal. Finally, the remaining ICs were visually inspected to further distinguish brain from non-brain (e.g., muscle) ICs based on each IC's equivalent dipole location, scalp topography, power spectrum, and power-power coupling estimated with the PowPowCAT toolbox,^36^ and non-brain ICs were removed. The final IC solution was then projected back onto the BeMoBIL preprocessed continuous channel data without the high-pass filter, because high-pass filters above 1 Hz, which improve AMICA decomposition, can distort event-related potential (ERP) waveforms.^37,38^ The cleaned data were then low-pass filtered with a cutoff frequency of 30 Hz, epoched from -2 to 2 seconds, and baseline subtracted from -150 to -50 ms before perturbation onset.^7,33^

Cortical N1 responses

Cortical N1 responses were extracted from the midline frontocentral electrode (Cz). To identify the N1 consistently across conditions for each individual, we first averaged all forward perturbation trials to create a composite waveform for each participant. The reference N1 latency was identified as the most negative local minimum within a 100–300 ms post-perturbation window; 2 participants (1 stroke, 1 control) required an extended time window (100-350ms) and 6 participants (3 stroke, 3 control) necessitated a shorter time window (100-250ms) in order to reliably capture the first local minimum in the cortical response waveform across all balance conditions. Next, the ERP waveforms were plotted separately for each condition of interest (Small-FIP, ST-step, ST-FIP). The N1 peak for each distinct condition was identified as the local minimum closest in latency to the N1 latency from the composite waveform across all trials. This sequence was performed automatically using MATLAB and visually confirmed for accuracy. N1 peak amplitudes were sign-inverted such that larger positive values denote larger cortical responses. N1 scaling (∆N1 amplitude) was quantified as the percent change in mean N1 peak amplitude and latency from the Small-FIP to the ST-FIP condition [100% × (ST-FIP − Small-FIP) / Small-FIP]. The equivalent percent change from the ST-FIP to the ST-step condition was computed to index the influence of behavioral outcome at the same ST perturbation magnitude. We also quantified the raw change in N1 amplitude (µV) and latency (ms) between these conditions.

### Statistical analysis

Normality and heterogeneity of variances were assessed using the Shapiro-Wilk and Levene’s test, respectively. Independent t-tests and Mann-Whitney U tests were conducted to examine group-level differences on normally-distributed and non-normally distributed metrics, respectively. The proportion of participants in each group showing an increase in N1 amplitude with perturbation magnitude was compared using Fisher's exact test. Two multiple linear regression models were constructed to determine whether stroke status moderated the association between N1 scaling and (1) N1 amplitude in the Small-FIP condition and (2) step threshold performance. Each model included group, the predictor of interest, and their interaction. Because relationships between cortical N1 responses and balance and mobility function have been shown to differ between clinical and nonclinical populations,^7,12^ simple slopes were estimated within each group a priori, regardless of the significance of the interaction term. Where the interaction was not significant, the model was refit excluding the interaction term to estimate the association across both groups. Coefficients are reported as standardized beta values. Multicollinearity was evaluated using predictor-level generalized variance inflation factors (all GVIFs ≤ 1.1). Linear mixed model analyses were conducted to examine whether the effects of perturbation magnitude (Small-FIP, ST-FIP) and step threshold behavioral outcome (ST-FIP, ST-step) on N1 amplitude and latency differed within and between groups. Post hoc analyses were conducted for significant interactions using pairwise comparisons with Bonferroni corrections. The assumptions of normality, linearity, and homoscedasticity were verified via residual plots. All analyses were performed using R software version 4.5.1 with a priori level of significance set to 0.05.

*Reference numbers correspond to the main manuscript reference list.*
